# A Novel Perspective on Genes Driving Metastatic Pancreatic Cancer Revealed by Single-cell RNA Sequencing

**DOI:** 10.1101/2024.03.03.24303664

**Authors:** Saed Sayad, Mark Hiatt, Hazem Mustafa

## Abstract

**Background:** Pancreatic ductal adenocarcinoma (PDAC) is formidable in its advanced, metastatic stage. Aggressive spread of malignant cells from the pancreas to distant organs like the liver and lungs is often detected late, complicating treatment by markedly limiting therapeutic options and worsening prognosis by drastically diminishing survival. Understanding the molecular intricacies driving metastasis is crucial for developing targeted therapies for this deadly disease with otherwise narrow recourse.

**Method:** We obtained single-cell transcriptomes (*GSE154778*) from the website of the United States National Institutes of Health. The single-cell RNA profiles of 10 PDAC primary tumors and six metastatic lesions, dissociated from one another, were obtained using the 10x Genomics Chromium platform. Our analysis focused on identifying genes, pathways, and gene ontology terms with distinct expression patterns between metastatic and primary single cells.

**Results:** Through single-cell RNA-sequencing (RNA-seq), we discerned significant alterations in gene expression profiles between primary tumors and metastatic lesions in PDAC, particularly emphasizing the dysregulation of ribosomal protein (RP) gene family as potential drivers of aggressive cancer behavior. Moreover, the enrichment of pathways related to metabolism, hypoxia response, and microbial influences underscores the intricate interplay between cellular adaptations and the tumor microenvironment in facilitating metastasis. Conversely, the downregulation of signaling pathways and extracellular matrix remodeling suggests a loss of regulatory control and enhanced invasive potential in metastatic cells.

**Conclusions:** In our comparison of primary and metastatic PDAC using single-cell RNA-seq, we have identified numerous differentially expressed genes, pathways, and gene ontology terms. The most significant finding may be that the ribosomal protein (RP) gene family is shared by 48 of the top 50 overexpressed pathways (comprising 5,848 genes), meaning that altering any member of this family as a potential driver could affect 48 pathways simultaneously. This revelation that metastatic cells may be regressed to a non-metastatic state by downregulating the RP gene family presents a promising pathway since this family is druggable.

## Introduction

Metastatic pancreatic ductal adenocarcinoma (PDAC) represents an advanced and often devastating disease, characterized by the spread of malignant cells from the pancreas to distant organs and tissues. Pancreatic cancer itself is notorious for its aggressive nature and often evades detection until it reaches an advanced stage, making treatment challenging and prognosis poor. However, when the tumor metastasizes, typically to organs such as the liver, lungs, and peritoneum, the complexity of managing the disease increases markedly. Metastatic spread not only diminishes treatment options, but also significantly reduces the likelihood of a successful outcome. The prognosis for patients with metastatic PDAC remains particularly grim, with a five-year survival rate of only 3% (1). In this study, we delve into genomic and molecular profiling, with a particular focus on the transformative potential of single-cell RNA-sequencing (RNA-seq). By harnessing the power of this cutting-edge technology, we have uncovered the pathways intricately involved in the metastasis of PDAC. This revelation not only deepens our understanding of the disease, but also paves the way for developing targeted interventions. Through meticulous analysis and interpretation of the data generated, we aim to contribute to ongoing efforts to combat this overwhelming illness.

### Data

We downloaded single-cell transcriptomes (*GSE154778*) from the website of the National Institute of Health. The single-cell RNA profiles of 10 PDAC primary tumors and six metastatic lesions, dissociated from one another, were obtained using the 10x Genomics Chromium platform (**Figure 1**).

**Figure 1:**
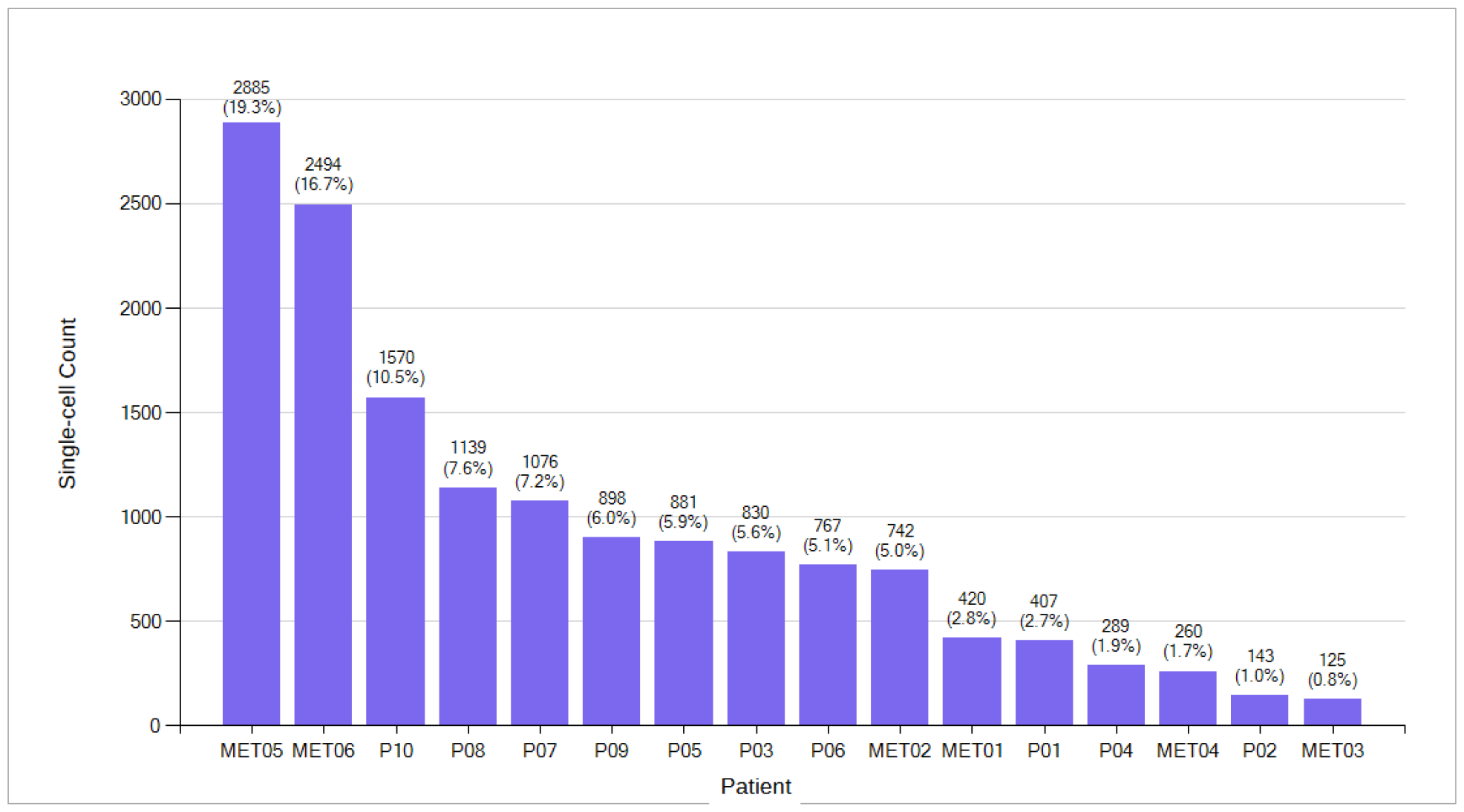
Single-cell count for 10 primary (P) and six metastatic (MET) PDAC patients.

### Data Analysis

In this study, we conducted a comprehensive comparative analysis of single-cell transcriptomes derived from both primary and metastatic PDAC. Our aim was to understand how gene activity changes as cancer progresses. Studying single-cell genetic data enabled us to look closely at each cell, helping us spot small differences in how genes were functioning. In this manner, we learned about the different types of cells involved in pancreatic cancer and how they changed over time. Since the primary tumor is the epicenter of malignancy, understanding the transcriptomic alterations occurring at this locale is pivotal to unraveling the initial stages of cancer development. Furthermore, our investigation extended to metastatic lesions, providing a unique perspective on the molecular adaptations facilitating cancer cell dissemination to, and colonization of, distant sites. In our analysis, we employed the t-test for bivariate data and a false discovery rate threshold of less than 0.05 to minimize the risk of spurious findings.

#### Differential Gene Expression – Primary Cells compared to Metastatic Cells

We employed a t-test to analyze PDAC cells from 10 patients (8,000 cells) compared to metastatic cells from six patients (6,926 cells) in order to identify genes with differential expression (**Figure 2**). This analysis sheds light on the differences between primary and metastatic cancer cell populations, enriching our comprehension of pancreatic cancer dynamics.

**Figure 2:**
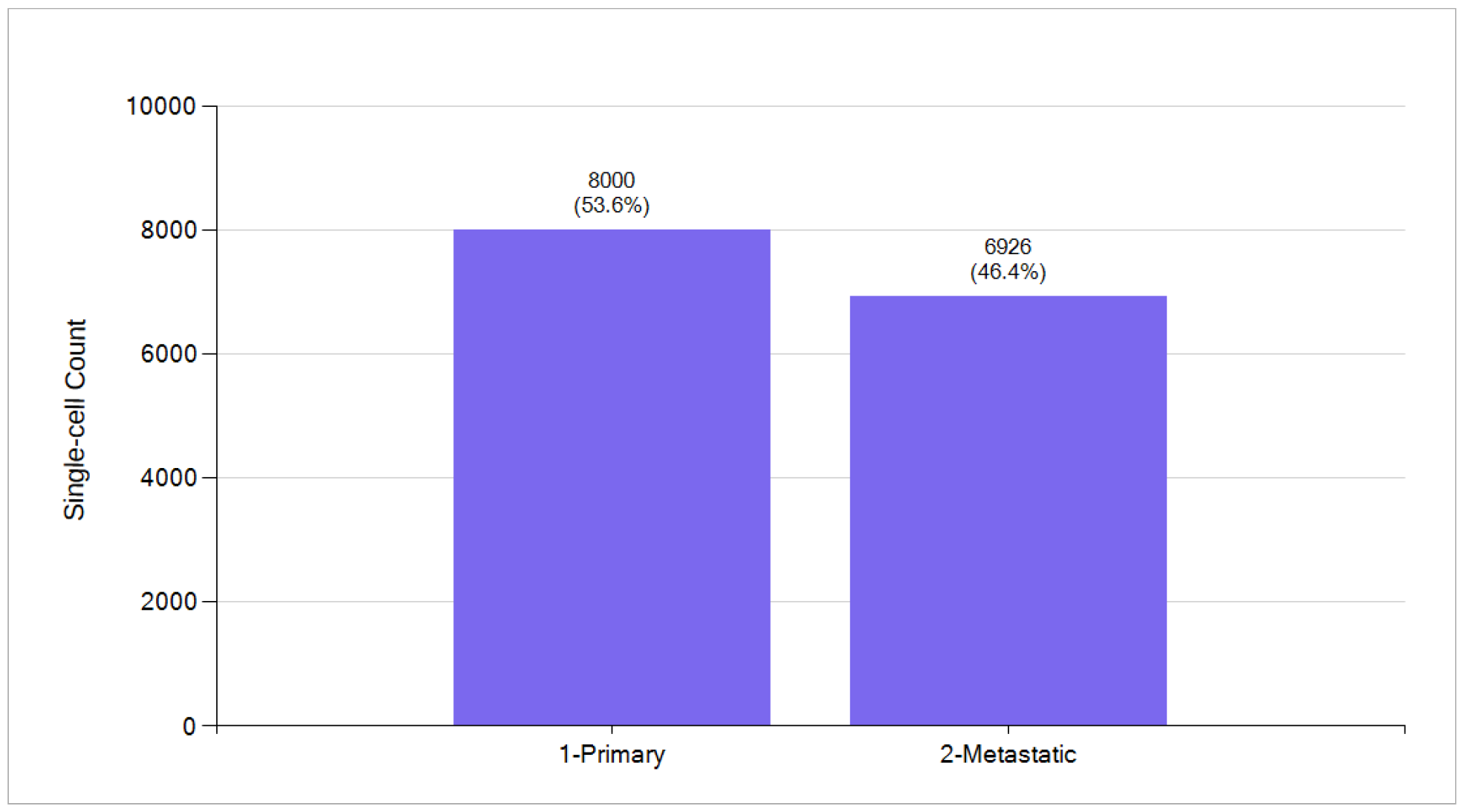
PDAC primary single cells from 10 patients and metastatic single cells from six patients.

From the numerous genes showing differential expression, we singled out the top 10 upregulated and downregulated genes (**Table 1**). Surprisingly, all the upregulated genes belong to the ribosomal protein (RP) gene family. Several RPs are overexpressed in various human tumors, including prostate, colon, hepatic, pancreatic, gastric, lung, and breast cancers, among others (2,3). Interestingly, nearly all the downregulated genes are associated with the *mitochondrially encoded NADH gene family* and *mitochondrially encoded cytochrome C and B* (4,5).

**Table 1:**
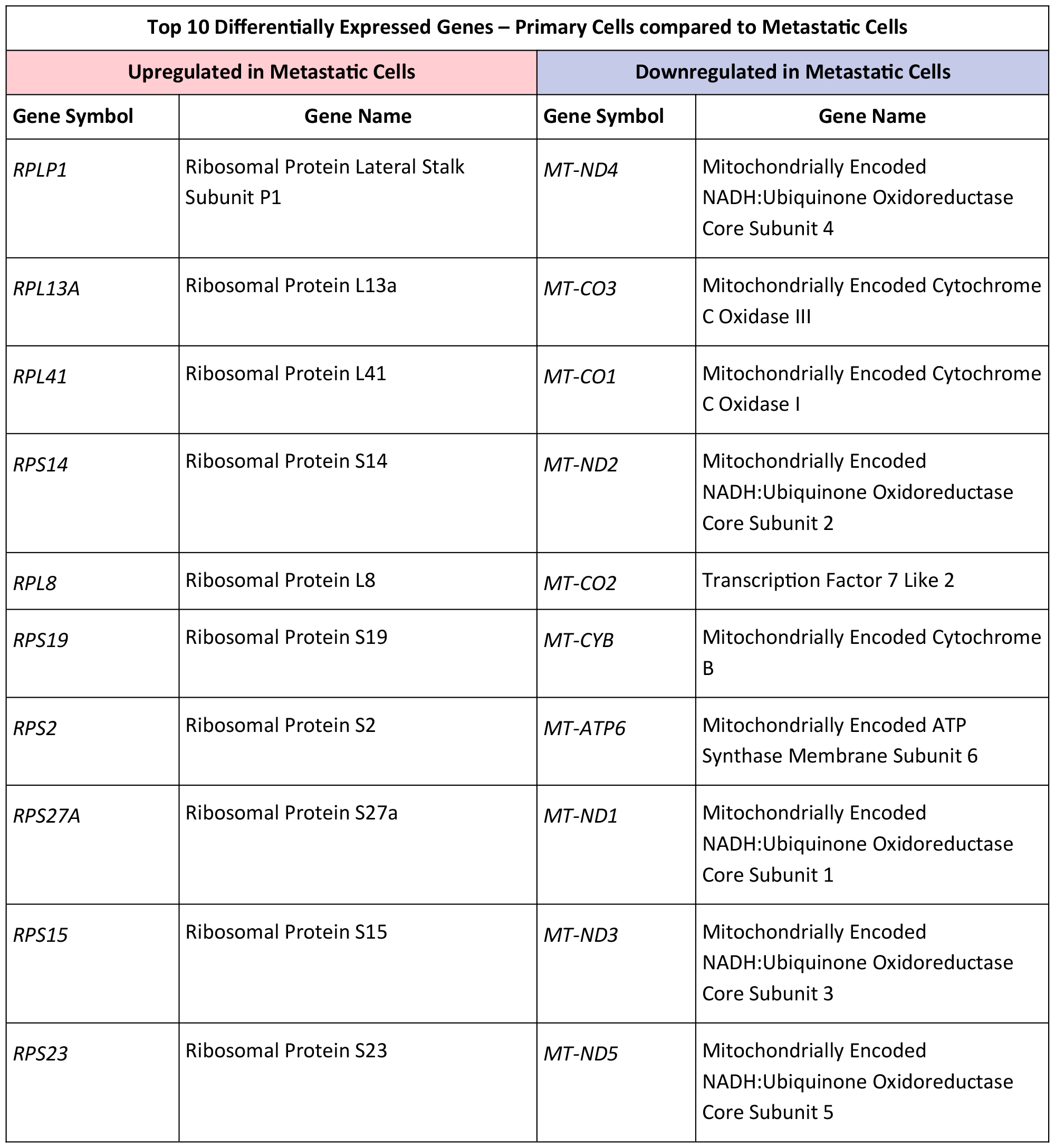
The top 10 upregulated and downregulated genes in metastatic cells compared to primary cells in PDAC.

In **Table 2**, metastatic cells display a range of upregulated pathways and mechanisms defined in the Kyoto Encyclopedia of Genes and Genomes (KEGG) that contribute to their invasive nature. Among these mechanisms are increased activity in *ribosome biogenesis*, facilitating enhanced protein synthesis necessary for metastasis (6). The *HIF-1 signaling pathway* is a major regulator of cellular response to changes in oxygen concentration, supporting the adaptation of tumor cells to hypoxia in an oxygen-deficient tumor microenvironment (7). Additionally, upregulation of *RNA transport* supports the efficient movement of genetic material within the cell, facilitating rapid adaptation to changing conditions (8). Metastatic cells also exhibit increased *thermogenesis*, potentially aiding in the energy-intensive processes associated with migration and invasion (9). The involvement of *proteoglycans* in cancer suggests a role in modulating cell-cell interactions and extracellular matrix (ECM) remodeling, further promoting metastatic behavior (10).

**Table 2:**
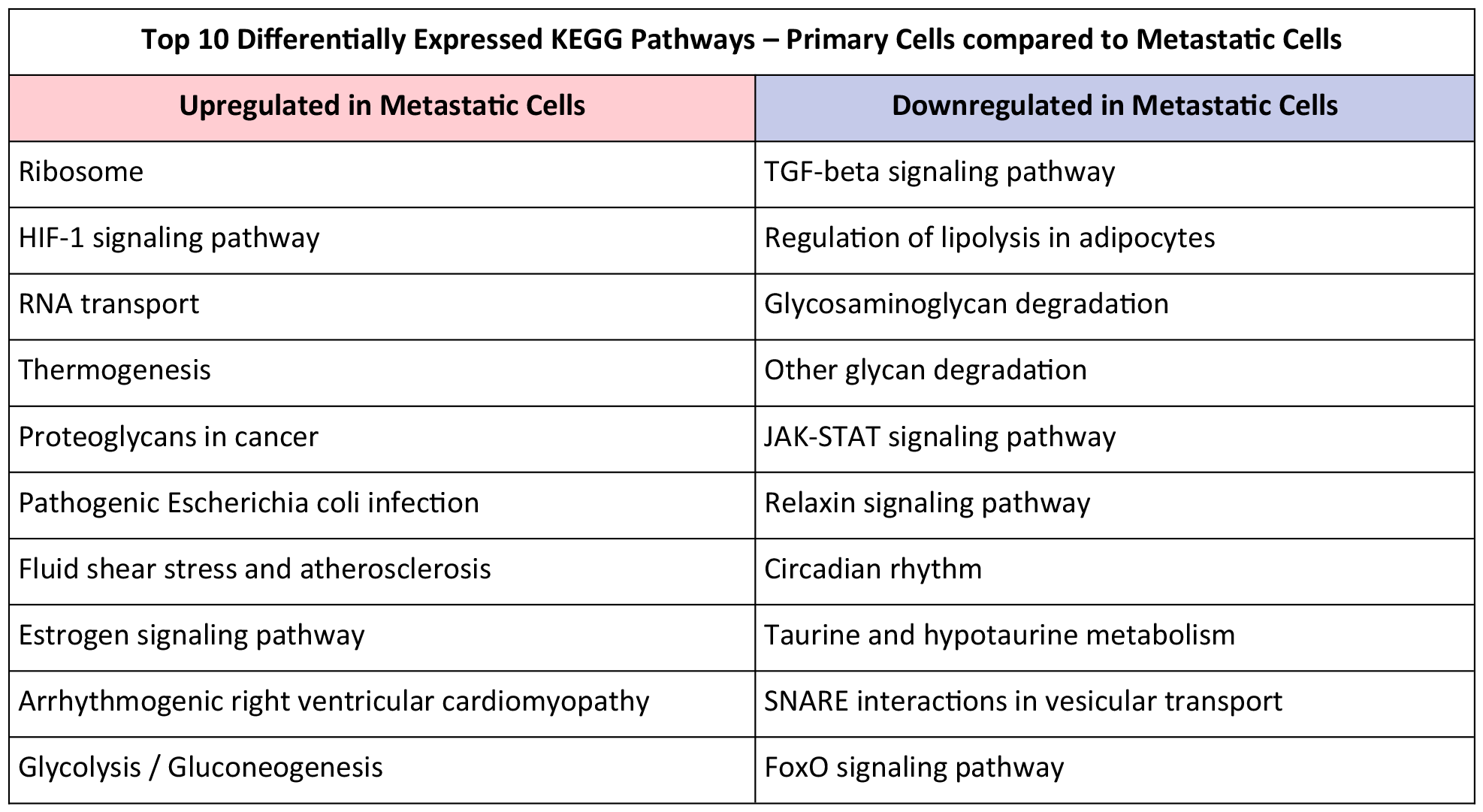
The top 10 upregulated and downregulated KEGG pathways in metastatic cells compared to primary cells in PDAC.

Furthermore, the presence of *pathogenic Escherichia coli infection* indicates a potential link between microbial factors and metastatic progression, highlighting the complex interplay between host and pathogen in cancer development (11). Additionally, *fluid shear stress* is implicated in atherosclerosis, suggesting a potential mechanism by which mechanical forces contribute to metastatic dissemination, particularly in vascularized tissues (12). The *estrogen signaling* pathway is also upregulated in metastatic cells, indicating hormonal influences on tumor progression, particularly in hormone-sensitive cancers (13). Finally, alterations in *glycolysis and gluconeogenesis* highlight metabolic reprogramming in metastatic cells, supporting increased energy demands and providing potential therapeutic targets for intervention (14). Overall, these upregulated pathways and mechanisms collectively contribute to the aggressive behavior of metastatic cells, highlighting the multifaceted nature of cancer progression.

In comparison to primary cells, metastatic cells exhibit downregulation of several key KEGG pathways and processes, as highlighted in **Table 2**. These include the *TGF-beta signaling pathway*, which plays a role in cell growth and differentiation and has been implicated in several hallmark features of PDAC pathobiology (15). *JAK-STAT signaling pathway* stimulates cell proliferation and malignant transformation and inhibits apoptosis in the pancreas (16). Various adipokines released from dysfunctional *adipocytes* have been reported to promote proliferation, invasion, metastasis, stemness, and chemoresistance of pancreatic cancer cells via different mechanisms (17). *Glycosaminoglycan* and *other glycan degradation* relate to the disruption of the major macromolecules composing the ECM, which spatiotemporally controls cell fate. Dysregulation of ECM remodeling can lead to tumorigenesis and cancer development by providing favorable conditions for tumor cells (18). Furthermore, suppressing *SNARE*s likely disrupts intracellular vesicle traffic, impairing invadopodium-associated protein transport. ECM modification, signaling pathway modulation critical for cancer cell movement (19), and suppressing *relaxin* likely exacerbate fibrosis and inflammation due to relaxin’s antifibrotic and anti-inflammatory properties (20). *Taurine and hypotaurine metabolism*, crucial for osmoregulation and antioxidation, is also affected (21). Finally, downregulation of *FoxO* can lead to dysregulated cellular processes, increased susceptibility to oxidative stress, impaired DNA repair, and enhanced tumorigenesis (22).

The main upregulated Reactome pathways in the metastatic pancreatic cancer cells (**Table 3**) are characterized by enhanced metabolism promoted by a hypoxic tumor microenvironment. Metabolic reprogramming allows cancer cells to survive hostile microenvironments (23). Additionally, recent studies have underscored the pivotal role of cellular stress in both initiating and sustaining cancer dormancy (24). The pivotal discovery reveals that the *RP gene family* is a common denominator in 48 of the top 50 upregulated Reactome pathways, encompassing 5,848 genes, implying that modifying one or more of these genes as potential drivers could impact 48 pathways concurrently. From this revelation, the possibility arises that metastatic cells may be reversed to a non-metastatic state by suppressing the RP gene family.

**Table 3:** The top 10 upregulated and downregulated Reactome pathways in metastatic cells compared to primary cells in PDAC.

| Top 10 Differentially Expressed Reactome Pathways – Primary Cells compared to Metastatic Cells |  |
| --- | --- |
| Upregulated in Metastatic Cells | Downregulated in Metastatic Cells |
| Metabolism of proteins | CS/DS degradation |
| Metabolism | Crosslinking of collagen fibrils |
| Disease | Defective ST3GAL3 causes MCT12 and EIEE15 |
| Developmental Biology | Defective CHST6 causes MCDC1 |
| Infectious disease | Keratan sulfate degradation |
| Metabolism of RNA | Scavenging by Class H Receptors |
| Cellular responses to external stimuli | Diseases associated with glycosaminoglycan metabolism |
| Cellular responses to stress | Dermatan sulfate biosynthesis |
| Nervous system development | Defective CHSY1 causes TPBS |
| Axon guidance | ECM proteoglycans |

The majority of downregulated Reactome pathways (**Table 3**) are related to the ECM, which plays a crucial role in PDAC, influencing its progression and affecting the effectiveness of treatments against it. By governing processes like migration, proliferation, anti-apoptosis, and cell metabolism, the ECM significantly contributes to shaping the cancer hallmarks in pancreatic cancer (25). For example, chondroitin sulfate and dermatan sulfate (CS/DS) can bind to various cytokines and growth factors, cell surface receptors, adhesion molecules, enzymes, and fibrillar glycoproteins of the ECM, thereby influencing both cell behavior and the biomechanical and biochemical properties of the matrix, highlighting the importance of CS/DS metabolism in cancers (26). Scavenger receptors, known for their diverse roles in various physiological and pathological conditions such as lipid metabolism, inflammation, and autoimmune disorders, are increasingly recognized as crucial regulators in tumorigenesis, cancer invasion, and the antitumor immune response, although their precise mechanisms and extent of involvement in cancer biology and immunology remain relatively unexplored (27).

Analysis of upregulated gene ontology terms in metastatic cells compared to primary cancer cells in PDAC reveals several key biological processes (BP) and cellular components (CC) implicated in metastasis (**Table 4**). *Exosomes* are extracellular vesicles that play a crucial role in the progression, metastasis, and chemoresistance of pancreatic cancer (28). Among the cellular components, *membrane* (29), *focal adhesion* (30), *cytosol, nucleolus*, and *endoplasmic reticulum* are notably upregulated (31). These components are associated with diverse cellular functions, including RNA binding, rRNA processing, and regulation of translation, suggesting a potential role for dysregulated gene expression and protein synthesis in the metastatic phenotype.

**Table 4:** The top 10 upregulated and downregulated gene ontology terms in metastatic cells compared to primary cells in PDAC.

| Top 10 Differentially Expressed Gene Ontology Terms – Primary Cells compared to Metastatic Cells |  |
| --- | --- |
| Upregulated in Metastatic Cells | Downregulated in Metastatic Cells |
| (CC) extracellular exosome | (CC) respiratory chain complex I |
| (MF) RNA binding | (MF) oxidoreduction-driven active transmembrane transporter activity |
| (CC) membrane | (BP) ATP synthesis coupled electron transport |
| (CC) focal adhesion | (CC) proton-transporting ATP synthase complex |
| (CC) cytosol | (CC) proton-transporting ATP synthase complex, coupling factor F(o) |
| (CC) nucleolus | (BP) ATP synthesis coupled proton transport |
| (BP) rRNA processing | (BP) electron transport coupled proton transport |
| (BP) regulation of translation | (BP) response to light intensity |
| (BP) negative regulation of translation | (MF) proton-transporting ATP synthase activity, rotational mechanism |
| (CC) endoplasmic reticulum | (BP) hyperosmotic salinity response |

The list outlines downregulated gene ontology terms in metastatic cancer cells compared to primary cancer cells in PDAC, indicating potential functional alterations associated with metastasis (**Table 4**). These cells exhibit decreased activity in several crucial biological processes (BP) and molecular functions (MF). Notably, downregulation occurs in *respiratory chain complex I* and *proton-transporting ATP synthase complexes*, suggesting compromised energy metabolism (32). Additionally, components related to energy production and transport are decreased, such as *ATP synthesis coupled electron transport* and *ATP synthesis coupled proton transport*. The reduction in *proton-transporting ATP synthase* activity, specifically in its rotational mechanism, underscores alterations in mitochondrial function, which could contribute to the metastatic phenotype (33). The decrease in *oxidoreduction-driven active transmembrane transporter* activity further indicates disruptions in cellular transport processes. Moreover, responses to environmental stimuli like *light intensity* and *hyperosmotic salinity* are also dampened, hinting at broader regulatory changes (34).

## Discussion

PDAC represents a daunting challenge in oncology, characterized by its aggressive nature and dismal prognosis. Our analysis of single-cell transcriptomes from PDAC primary tumors and metastatic lesion biopsies provided valuable insights into the molecular alterations associated with cancer progression. By comparing gene expression profiles between primary and metastatic cells, we identified significant changes in key pathways and biological processes implicated in metastasis. Notably, our findings shed light on the dysregulation of the RP gene family in metastatic cells, highlighting their potential role as drivers of aggressive cancer behavior. The upregulation of members of this family across multiple pathways underscores their central importance in orchestrating cellular processes conducive to metastasis. Furthermore, our analysis revealed the enrichment of pathways related to metabolism, hypoxia response, RNA transport, and proteoglycan signaling in metastatic cells. These findings suggest a metabolic reprogramming and adaptive response to the tumor microenvironment, facilitating the survival and dissemination of cancer cells. Additionally, the identification of pathogenic *E. coli* infection and estrogen signaling pathway activation in metastatic cells highlights the complex interplay between microbial factors, hormonal influences, and cancer progression. In contrast, metastatic cells exhibited downregulation of pathways associated with cellular signaling, ECM remodeling, and cellular stress response. The suppression of key pathways such as TGF-beta signaling, JAK-STAT signaling, and ECM remodeling suggests a loss of regulatory control and dysregulated interactions with the tumor microenvironment. These alterations may contribute to the acquisition of invasive phenotypes and resistance to therapeutic interventions.

Our analysis also showed changes in gene ontology terms associated with cellular components and biological processes in metastatic cells. Upregulated components such as exosomes, focal adhesions, and cytosolic machinery reflect the dynamic changes in cellular architecture and function associated with metastasis. Conversely, the downregulation of components involved in energy metabolism, cellular transport, and environmental response pathways highlights the metabolic and functional alterations accompanying the metastatic transition. Overall, our study provides insights into the genomic and molecular landscape of metastatic pancreatic cancer, uncovering potential therapeutic targets and pathways for intervention. By elucidating the intricate mechanisms driving metastasis, we aim to contribute to the development of novel treatment strategies and personalized therapeutic approaches for patients facing this devastating illness. However, further studies are warranted to validate our findings and translate them into clinical applications, ultimately improving outcomes for patients with metastatic pancreatic cancer.

## Summary

Metastatic PDAC presents a formidable challenge in oncology due to its aggressive nature and dismal prognosis. Using single-cell RNA-sequencing our analysis revealed numerous differentially expressed genes, pathways, and gene ontology terms and the significant finding that the RP gene family is shared among 48 of the top 50 overexpressed pathways (comprising 5,848 genes), meaning that altering a RP gene as a potential driver could affect 48 pathways simultaneously. Such alteration affecting the downregulation of this family, given its druggable nature, may be a feasible means to regress metastatic cells to a non-metastatic state.

## Data Availability

All data produced are available online at:
https://www.ncbi.nlm.nih.gov/geo/query/acc.cgi?acc=GSE154778

